# Trends in opioid seizure data and their association with opioid mortality

**DOI:** 10.1101/2022.10.08.22280845

**Authors:** Heather McBrien, Monica Alexander

## Abstract

**Background:** The opioid epidemic remains an emergent health issue in the United States, as opioid-related deaths continue to rise in the second year of COVID-19. The introduction of synthetic opioids into the illicit supply began causing deaths in 2015, however, data describing the illicit opioid supply is scarce.

**Methods:** We used a newly available national dataset of drug seizure reports, aggregated from law enforcement agencies across the United States, to describe changes in fentanyl, heroin, and other opioid presence in the national illicit supply from 2011-2017, by state and geographic region. We assessed the relationship between drug seizures and opioid-related deaths at the state level using linear regression.

**Results:** National and state increases in opioid seizure rates from 2011-2017 were entirely due to increased fentanyl and heroin seizures, as other opioid seizure rates remained constant. Most increases in seizures occurred in the Northeast, Midwest, and Appalachia, where fentanyl seizures and heroin seizures were highest and increased most sharply along with opioid deaths. The composition of drugs seized was similar within geographic regions, but did vary across regions. State opioid seizures of all types were strongly associated with state opioid deaths. The strongest relationship was between fentanyl seizures and fentanyl deaths.

**Conclusions:** The association between opioid seizures and deaths means seizure data has potential as an early-warning system to predict overdose, although national level data requires quality improvement. Regional variation in seizure rates supports existing evidence that illicit fentanyl and heroin supplies differ between regions, producing distinct regional risk environments, causing varying mortality rates.

## 1 Introduction

The opioid epidemic remains an emergent public health issue: 48 000 people died of opioid overdose in 2019 (Ahmad, Rossen, and Sutton 2020), and COVID-19 worsened opioid-related mortality, producing 20-year record drug overdose deaths in the 12 months before February 2021 (Woolf and Schoomaker 2019; Haley and Saitz 2020).

The three decade increase in opioid deaths can be divided into three waves, the first caused by opioid over-prescribing (Ciccarone 2019; Maxwell 2011; Kanouse and Compton 2015; Paulozzi et al. 2011), and made possible by aggressive marketing and availability of long-acting opioids (Dasgupta, Beletsky, and Ciccarone 2018). In contrast, changes in the illicit drug supply precipitated the second (heroin) and third (fentanyl) waves of the epidemic (Hedegaard, Minino, and Warner 2018; Kanouse and Compton 2015; Maxwell 2011; Mars, Rosenblum, and Ciccarone 2019). In 2011, heroin became cheaper and purer (Rosenblum, Unick, and Ciccarone 2014; Drug Enforcement Administration 2016), while prescription opioids became more regulated (Cicero et al. 2014; Beletsky and Davis 2017), broadening heroin use and heroin deaths (Trinidad et al. 2016; Shiels et al. 2019). Since the 2015 introduction of fentanyl into the illicit drug market (especially in the Northeastern US), opioid use is more risky and more likely to result in overdose (Zoorob 2019; O’Donnell et al. 2017; Hedegaard, Minino, and Warner 2018).

Valuable data such as administrative records (Bohnert et al. 2011; Miller et al. 2015), surveillance system data (Dart et al. 2015), clinical trials (Chou et al. 2015), and Medicaid data (Fulton-Kehoe et al. 2015) allow a detailed understanding of the first wave of the epidemic, and the causative relationship between excessive prescribing and deaths. However, unlike prescription opioids, illicit opioids produce no administrative data, and their illegality prevents any regulation, tracing, or data collection like that conducted with prescription opioids (Ruhm 2019; Ciccarone 2019; Fischer, Pang, and Jones 2020). Therefore, even though deaths caused by illicit opioids dominate the epidemic, the illicit drug supply changes likely causing these deaths are still mysterious (Ruhm 2019; Ciccarone 2019; Fischer, Pang, and Jones 2020).

Still, studies describe the arrival of fentanyl in the drug supply (Kenney et al. 2018), the adulteration of heroin with fentanyl (Macmadu et al. 2017; Carroll et al. 2017; Ciccarone, Ondocsin, and Mars 2017), and identify unwanted or accidental fentanyl exposure as causes of death (Mars, Ondocsin, and Ciccarone 2018; Ciccarone 2017). Reports from the DEA and independent researchers have suggested that fentanyl-adulterated heroin is a significant driver of opioid mortality in the Northeast (Ciccarone 2017; Carroll et al. 2017; Kenney et al. 2018). Additionally, reports derived from forensic analysis of seized drugs and illicit drug research programs corroborate the narratives of fentanyl introduction into the drug supply alone and as an additive, (Drug Enforcement Administration 2017, 2020; National Forensic Laboratory Information System 2017). These studies suggest that opioid supply markets differ by geographic region, and that these differences produce varying regional overdose risks. However, surveys and qualitative research referred to above use small datasets, and the DEA and UN only share data from illicit drug research programs in the form of summary statistics (Drug Enforcement Administration 2020; Drugs and Crime 2012), meaning detailed, independent analysis of national illicit supply data is impossible.

A better understanding of when and where illicit opioids lead to deaths would allow for more effective prevention of drug overdose, because prevention resources could be allocated where and when people need them most. Recent work shows a strong relationship between drug seizures and opioid-related harm: Rosenblum et al., in their 2021 paper, investigated the potential of Ohio state crime lab data in describing changes in the illicit drug supply, while evaluating the relationship between illicit drug analyses and mortality (Rosenblum, Unick, and Ciccarone 2020a). They found Ohio county seizure data was strongly predictive of county opioid mortality, and following other authors (Zoorob 2019; Rosenblum, Unick, and Ciccarone 2020b), they suggested seizure data has potential as a surveillance tool to predict when overdoses may occur (Rosenblum, Unick, and Ciccarone 2020a). In 2019, Zoorob filed a Freedom of Information Act Request to access the DEA’s National Forensic Laboratory Information System (NFLIS), which collects the results of forensic analyses conducted on drugs seized as part of law enforcement investigations (National Forensic Laboratory Information System 2020c). The forensic analyses in the NFLIS dataset represent 98% of all analyses by law enforcement investigations carried out in the US since 2010, and the dataset spans 50 states and 50 state lab systems (National Forensic Laboratory Information System 2020a). Zoorob showed that fentanyl seizures were strongly predictive of fentanyl deaths by state, and again called for the use of seizure data in an early-warning surveillance system.

In this paper we expand on this work, using the same NFLIS dataset to describe and quantify opioid seizures in the United States, and assess the correlation between opioid seizures and opioid mortality. We aim to:

1. Describe trends in the NFLIS opioid seizure data by year and state and by year and region.
2. Describe changes in the relative prevalence of heroin, fentanyl, and non-heroin-non-fentanyl opioid seizures, and heroin and fentanyl seized together, by year and state, and by year and region.
3. Evaluate the strength of association between NFLIS drug seizure reports and drug-related mortality.

We hope to better understand how illicit opioids lead to deaths through a close examination of this dataset.

## 2 Data

The National Forensic Laboratory Information Reporting System (NFLIS) aggregates lab results from forensic analyses by law enforcement agencies such as the DEA, customs and border agencies, and local police departments, producing one national dataset. Data entries are results from lab analyses of seized drugs, analyses of undercover drug buys, or analyses of other evidence in law enforcement investigations. Private forensic labs or state lab systems report results voluntarily. Though there are private forensic labs used by law enforcement that do not participate, according to NFLIS, the dataset covers 98% of all forensic analyses in the United States (National Forensic Laboratory Information System 2020a), with 283 participating labs and 50 state lab systems, and has maintained similar coverage since 2011.

Data reports to NFLIS are different between states: 22 states report test-level data, where each row details a single analysis, in which up to eight drugs may have been identified. Other states report the total number of analyses containing a combination of drugs each month [^1]. NFLIS publishes yearly reports on the number and type of drug seizures they register, available at [linked phrase] https://www.nflis.deadiversion.usdoj.gov/Reports.aspx. NFLIS reports ∼1.5 million analyses per year. Over the entire study period, we analyzed just under 10 million reports.

The time between law enforcement drug acquisition and lab analysis also varies by state and by lab. NFLIS receives data for inclusion from a certain ‘period of interest’ within three months of the end date of the period. Around 5% of cases are submitted after this period, and are not included. Additionally, NFLIS reports that 30% of drug seizures nationally are never analyzed (National Forensic Laboratory Information System 2020b). Missingness from late reporting is therefore small, but missingness from never-analysed samples is significant. We conducted our analysis at the yearly level using the date of analysis to minimize the effect of delayed reporting, and smooth other sources of time-varying error.

Supplementary information on drug purity, the quantity of drug present in a report, and information on how drugs were acquired by law enforcement (ex. seized, purchased during policing, free sample, etc.) is included in the NFLIS data, but not reported uniformly between states. Where states report the total number of drugs seized as a single summary report, average purity measurements and the total sum quantity of drug seized are given. Purity and quantity information is missing in most summary reports – 86% and 40% respectively – and in some individual level reports – 96% and 33% respectively. Information on mode of acquisition is 84% missing.

Due to poor reporting, we did not comment on drug purity, nor did we use acquisition information to distinguish between reports, although the information which is present suggests that non-seizure reports are relatively rare, at 8%. We did not use drug quantity measurements either, but relied instead on number of reports as a measure of drug presence.

## 3 Methods

### 3.1 Description of national seizure rates by state and region

To describe opioid seizure trends by state and time, we developed a list of the 100 most frequent substances appearing in the dataset, and categorized these substances into 34 broad drug categories. A table of these categorizations is available in the appendix. These classifications covered 99.9% of the substances appearing the dataset, and ‘opioid’ was included in this list as a drug category. We could not manually categorize all drugs in the dataset because it contained millions of reports, nor could we search reports for opioids using keywords, as developing a comprehensive list of all opioids is impractical. We therefore matched substances listed in the dataset to our broad drug categories. We counted seizures containing at least one drug in the ‘opioid’ category as opioid seizures.

While identifying all opioid seizures by keyword is impractical, heroin and fentanyl are identifiable by name. To find heroin-containing seizures, we searched all substance fields in a report for “heroin” or “Heroin.” To identify fentanyl, we searched all fields in each report for any fentanyl or fentanyl analogue, using a list of drug names describing fentanyls from (Zoorob 2019) (see appendix). We included seizures containing any fentanyl or heroin in counts and plots of these drugs, even if there were multiple drugs reported in an analysis. To identify non-heroin-non-fentanyl opioid seizures, we removed all fentanyl or heroin-containing seizures (as defined above) from the list of opioid seizures.

To produce opioid seizure rates by state and year, we counted opioid seizures, fentanyl seizures, heroin seizures, and non-heroin-non-fentanyl seizures as defined above by state and year. We used data from the National Center of Health Statistics to estimate state-year populations. In the results section, we present descriptions of seizure rates by state and year, and discuss temporal and geographic trends in these estimates.

### 3.2 Relationship between seizures and mortality

To assess the relationship between opioid seizure rates and opioid mortality, we regressed fentanyl, heroin, and all opioid seizure rates (adjusted for population) against fentanyl, heroin, and all opioid mortality rates on the log scale. We used NCHS all-cause mortality data, identifying opioid-related deaths using the International Classification of Diseases, 10th Revision (ICD–10) underlying cause-of-death codes X40–X44, X60–X64, X85, and Y10–Y14. Because socioeconomic covariates such as median income and race, sex, or age structure did not change significantly over the study period, we relied on the state fixed effects to control for these differences, as well as differences in policing and reporting by state. We included year fixed effects for time-varying trends. Though we are primarily interested in the relationship between heroin and fentanyl seizures and mortality, because opioid overdose deaths are often under-reported or miscategorized, we also regressed all-opioids seizure rates against log opioid mortality, as this measure was likely to include all relevant deaths. We aimed only to evaluate whether there was any association between seizures and mortality, rather than to precisely model a hypothesized relationship.

We modeled the relationship between seizures and mortality to determine how well seizure data reflected the true illicit opioid supply. Strong evidence from the literature suggests that changes in illicit drug supply have caused overdose deaths, and therefore a positive association likely exists between true drug availability and mortality. If the NFLIS data reflected the true illicit drug supply, we expected to observe an association between seizures and mortality. A null result would have suggested data quality issues such as poor coverage, heterogeneous reporting between states, or changes in policing practices. It also may have suggested confounding: for example, law enforcement seizures may have increased during times of economic downturn, as policing practices change with funding, while at the same time overdoses increased due to the same economic changes.

## 4 Results and Discussion

### 4.1 Description of national seizure data

#### 4.1.1 State-specific seizures

Figure 1 shows opioid seizures per 100, 000 people by state during the study period. Seizure rates varied widely between states. Overall, they were higher in the Northeast and lower in the West. Absolute seizures were highest in Ohio, and increased throughout the study period. Massachusetts also reported high and increasing seizures, along with New Jersey and North Dakota.

**Figure 1:**
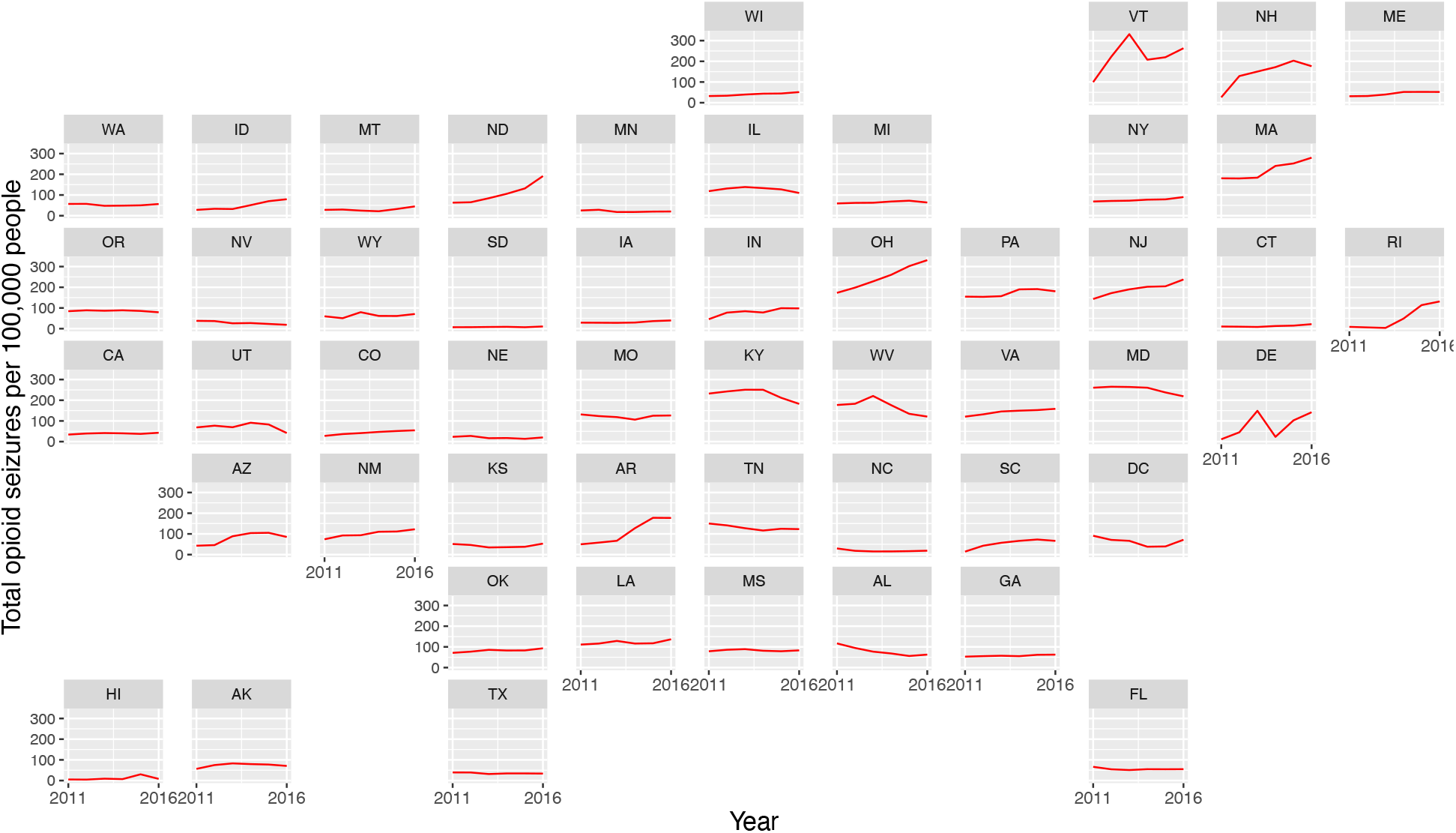
Standardized opioid seizure rates by state. Data from National Forensic Laboratory Information System and the National Center for Health Statistics.

Figure 2 shows heroin seizures per 100, 000 people by state, and fentanyl seizures per 100,000 people by state. Heroin seizures varied widely by region. They were orders of magnitude higher and varying in Northeastern states, but remained relatively constant in other areas of the country. New Jersey, Ohio, and Virginia all reported high and still increasing seizure rates throughout the study period, while heroin seizures appear to have peaked in several states including New Hampshire, Massachusetts, Pennsylvania, and Maryland.

**Figure 2:**
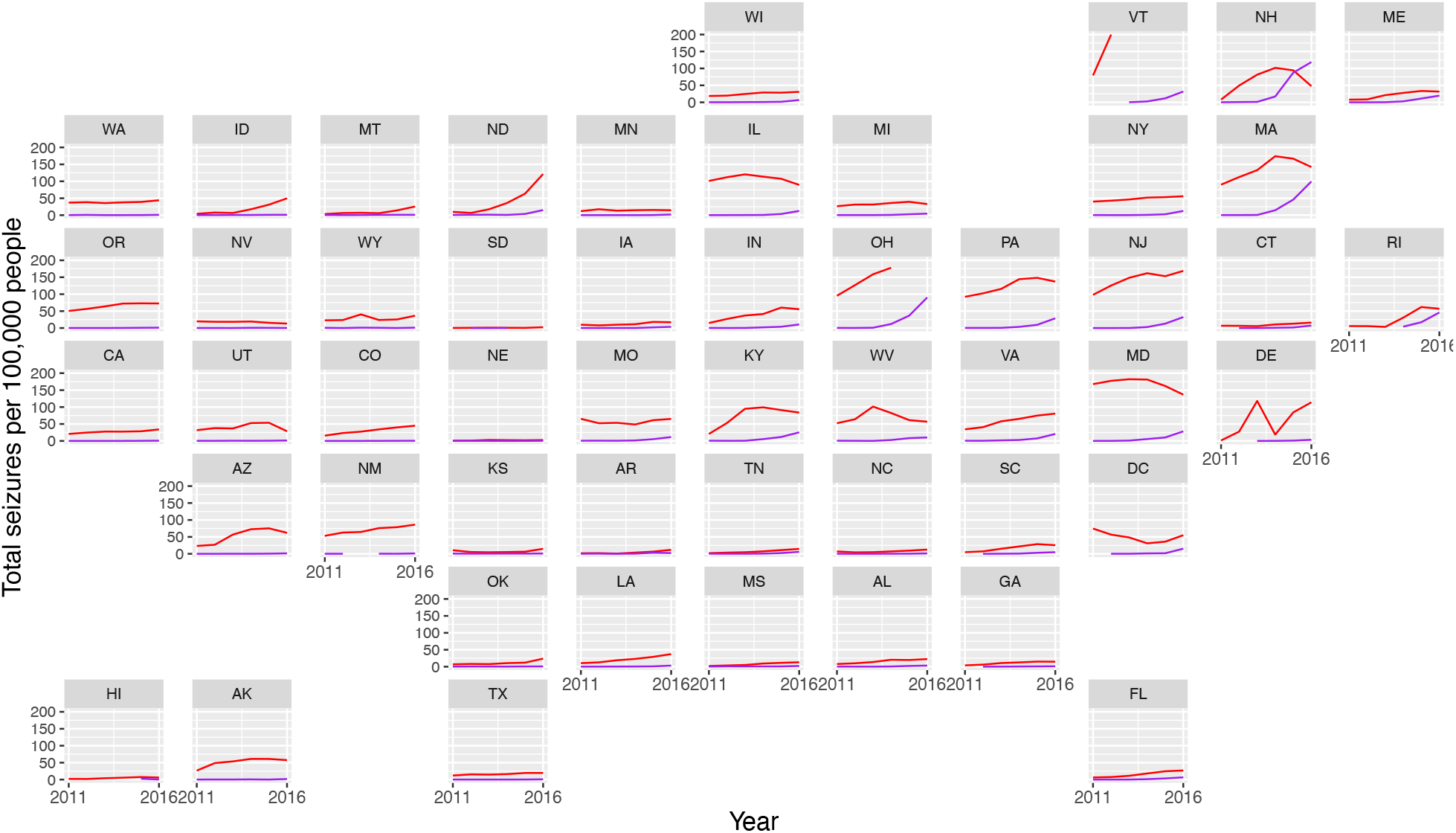
Standardized heroin and fentanyl seizure rates by state. Heroin is in red, fentanyl in purple. Data from National Forensic Laboratory Information System and National Center for Health Statistics.

There were also clear differences in fentanyl seizure rates across states. Like heroin seizures, fentanyl seizures remained relatively constant in Western and Southern states but increased exponentially from 2013 in Northeastern states. Small increases are visible in Western and Southern regions. Unlike heroin seizures, fentanyl seizure rates did not peak during the study period, and were high and increasing in Massacheusetts, Ohio, Pennsylvania, Virginia, Maryland, New Hampshire, and Vermont.

We categorized 99.9% of test result entries into 34 broad drug categories, of which ‘opioid’ was one. We identified non-heroin non-opioid seizures using this categorization, (referred to as ‘other opioid seizures’ from here), then excluded any seizures containing any fentanyl or heroin, for other opioid seizures, plotted in Figure 3. When considering only other opioid seizures, stark regional differences disappear, though state trends varied widely. Other opioid seizures decreased overall, and decreased in almost every state. Decreases were most pronounced in Kentucky, New Hampshire, Maryland, Massachusetts, and Illinois; all states with high opioid mortality. This pattern suggests that national increases in all opioid seizures, including fentanyl and heroin, are almost entirely attributable to increased heroin and fentanyl seizures.

**Figure 3.**
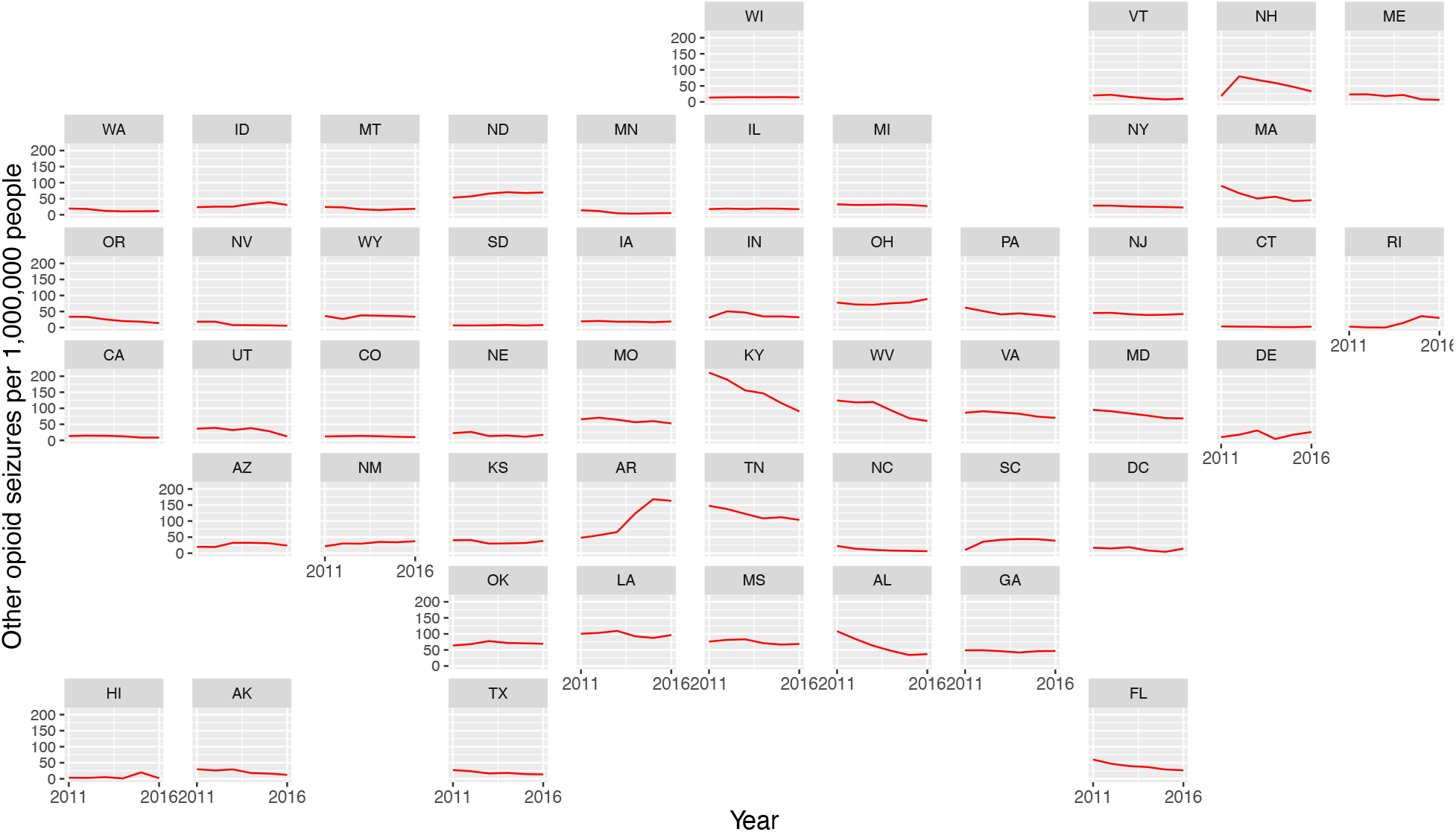
shows standardized seizure rates (seizures per 100,000 people) of non-fentanyl non-heroin opioids (‘other opioids’) by state.

#### 4.1.2 Regional seizure rates

While figures 1-3 illustrate differences in heroin and fentanyl seizure rates and trends over time, between states, they also show regional patterns. For example, fentanyl seizures increased over time in Eastern, Midwestern, and Appalachian states and remained relatively low in other states. These differences are summarized in Figures 4 and 5, which show seizure rates standardized by population in the Northeast, Appalachia, and Midwest as a single group and in the rest of the country. Small increases in fentanyl seizures were visible in regions other than this broad Northeast region; see Figure 5.

**Figure 4:**
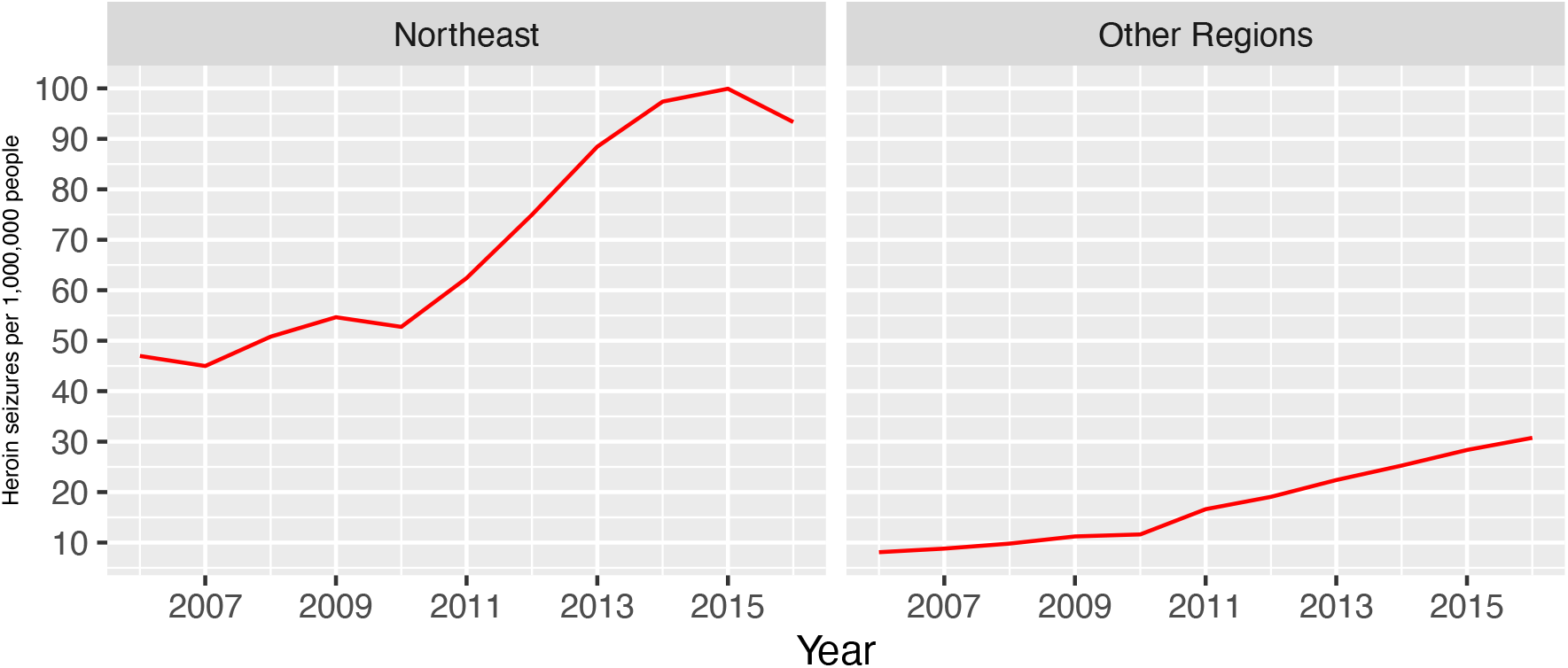
Standardized heroin seizures in the Northeast, Midwest, and Appalachia and standardized heroin seizures in the rest of the country. Data from National Forensic Laboratory Reporting System and National Center for Health Statistics.

**Figure 5:**
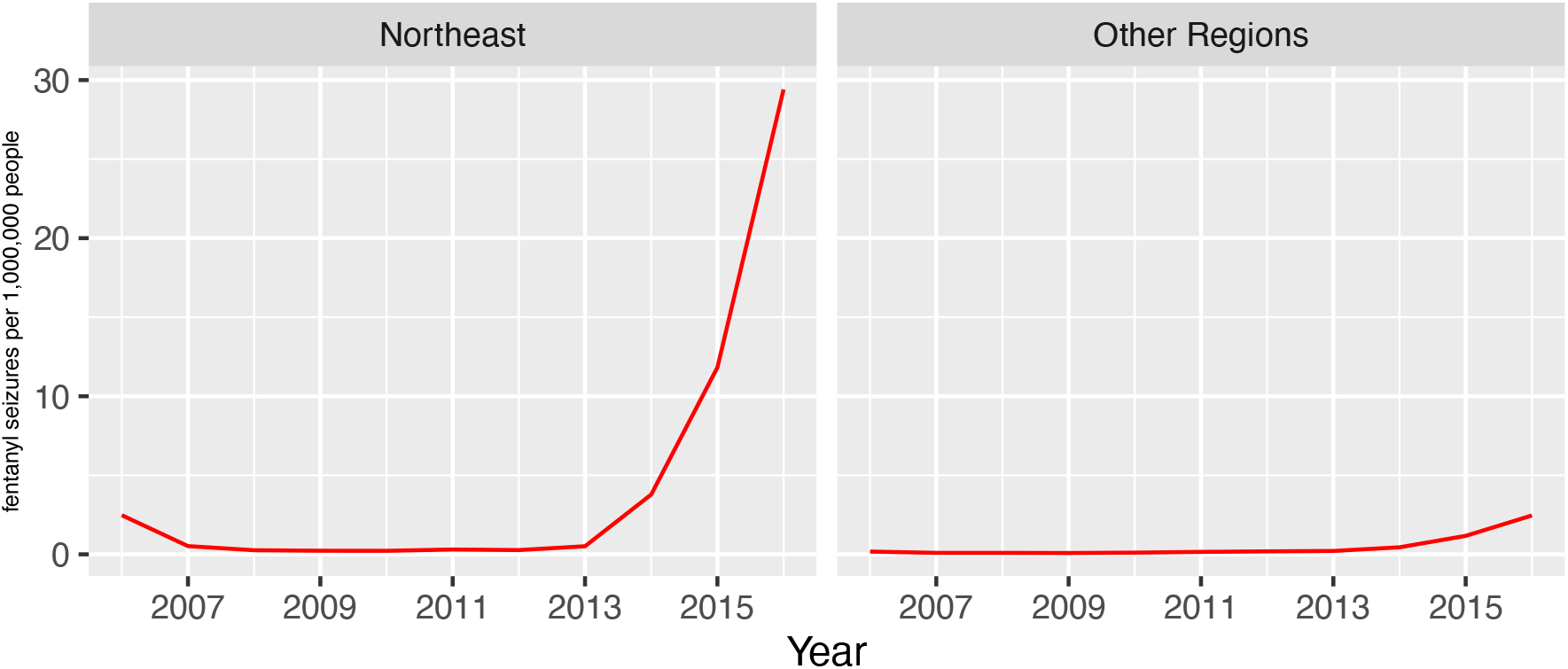
Standardized fentanyl seizures in the Northeast, Midwest, and Appalachia, and standardized heroin seizures in the rest of the country. Data from National Forensic Laboratory Reporting System and National Center for Health Statistics.

#### 4.1.3 Seizures of fentanyl and heroin together

Some studies have suggested that increases in fentanyl mortality are due to fentanyl-adulterated heroin in the Northeast. In particular, there have been qualitative reports of heroin containing fentanyl as an additive, and fentanyl being sold as heroin, leading to overdose (Ciccarone 2017; Carroll et al. 2017; Kenney et al. 2018). We looked for fentanyl and heroin seized together, which would support this narrative. Fentanyl and heroin seizures increased in some states. Both Ohio and Pennsylvania, two states with high fentanyl seizures and high fentanyl mortality, have seen dramatic increases in fentanyl and heroin seized together. Both have seen roughly a 2000% increase since 2011, plotted in Figure 6.

**Figure 6:**
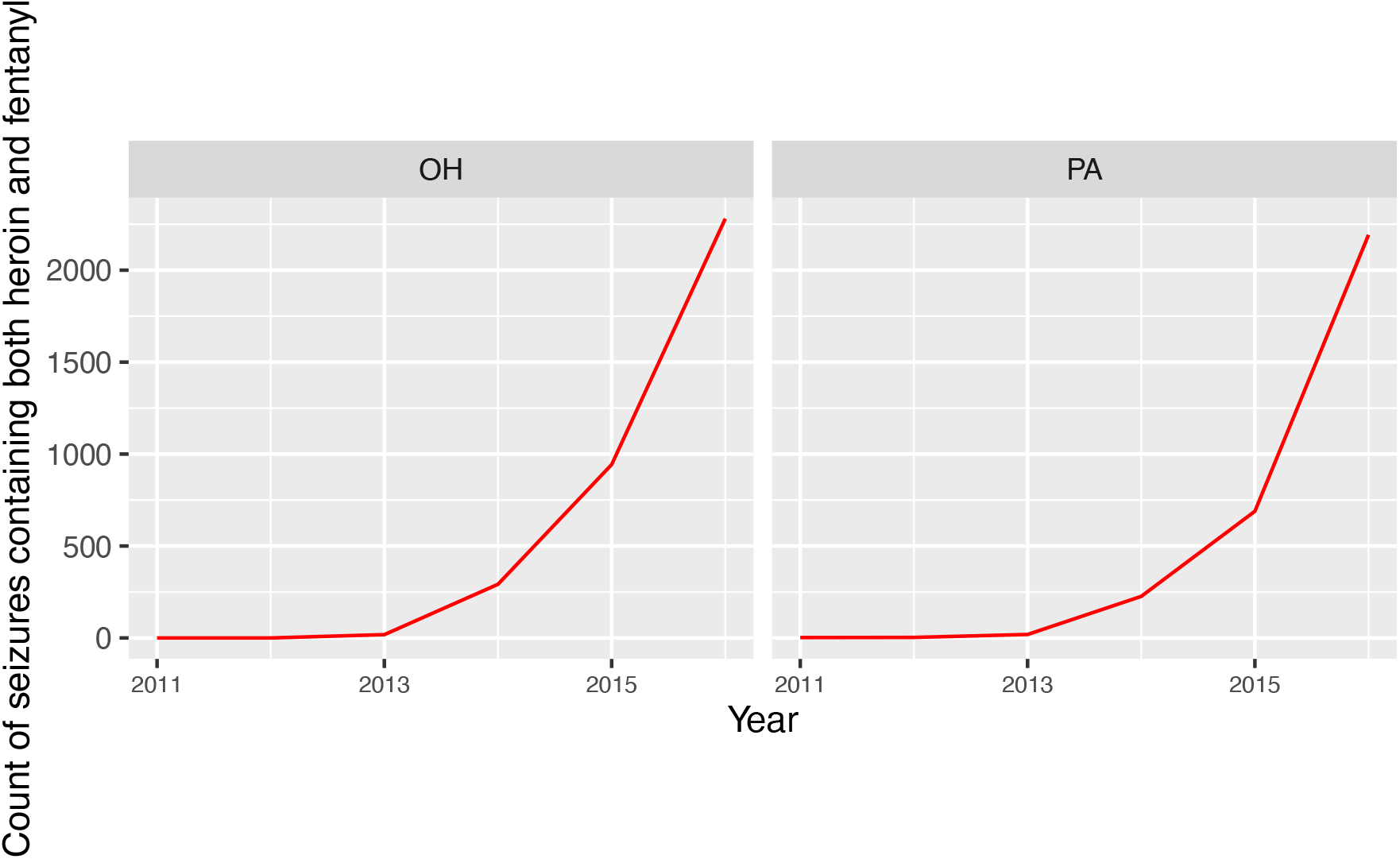
Counts of heroin and fentanyl seized together in Ohio and Pennsylvania. Data from the National Forensic Laboratory Information System.

### 4.2 Relationship between seizures and opioid mortality

Figure 7 shows relationships between opioid seizures per 100 000 people by year and state, and age-standardized opioid mortality rates by year and state, for all opioid seizures and all types of opioid mortality, fentanyl seizures and fentanyl-related mortality, and heroin seizures and heroin-related mortality.

**Figure 7:**
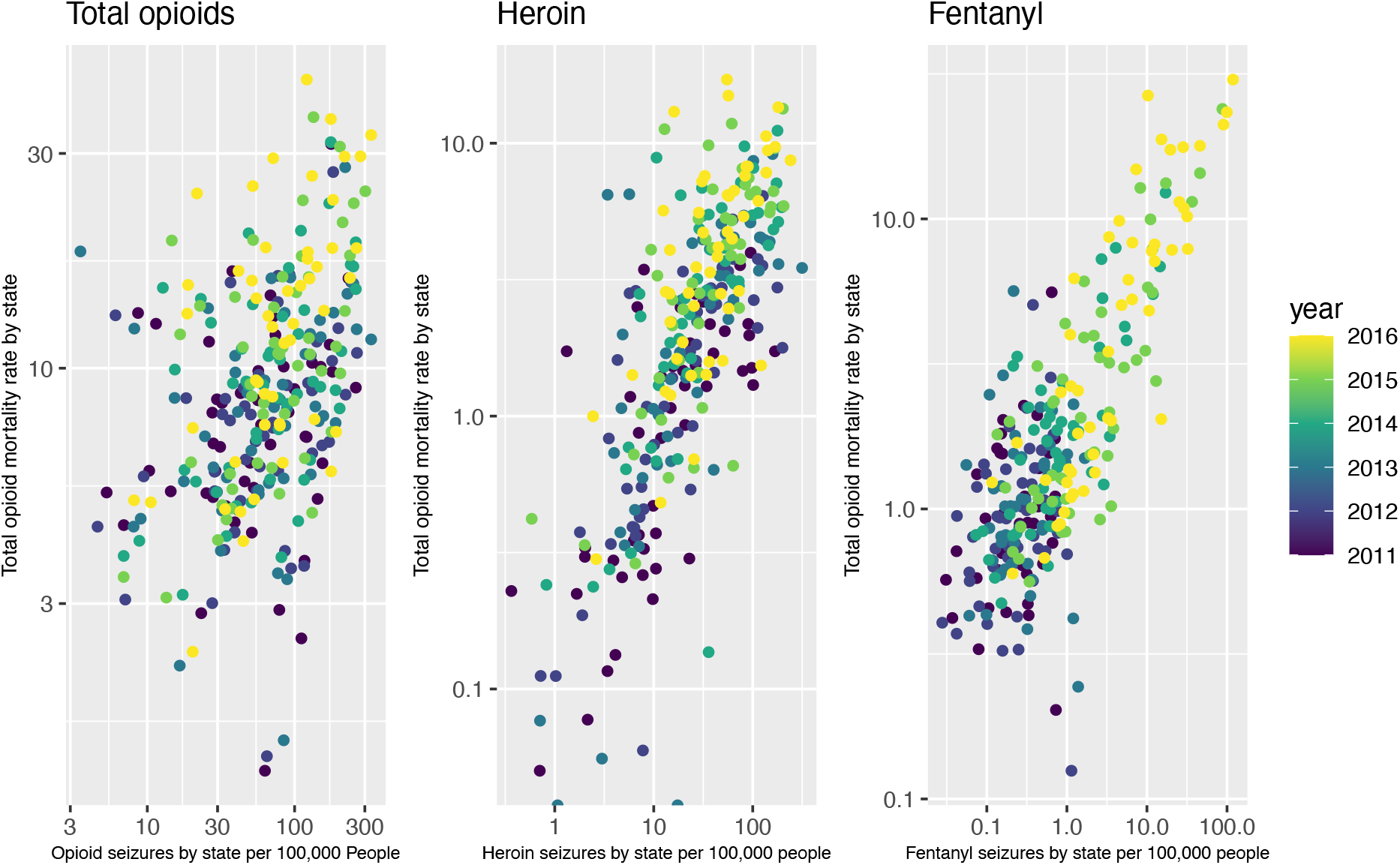
Scatterplot of standardized opioid seizure rates by year and state and opioid mortality rates by year and state for all opioid seizures and all types of opioid mortality, fentanyl seizures and fentanyl-related mortality, and heroin seizures and heroin-related mortality. Data from National Forensic Laboratory Reporting System and National Center for Health Statistics.

**Figure 8:**
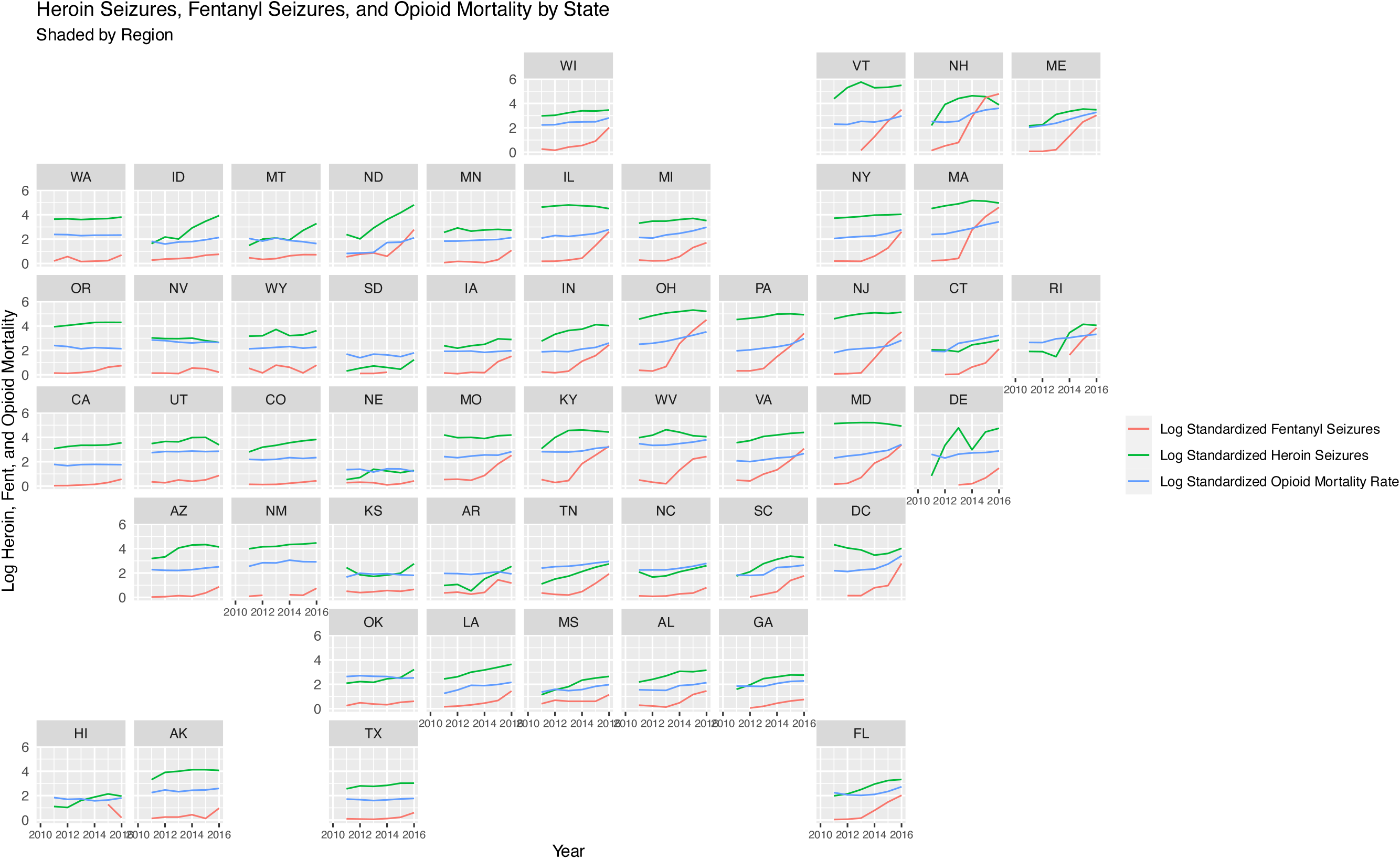
Opioid mortality rates and heroin and fentanyl seizure rates by year and state, coloured by region. Green states are Northeastern states with high and increasing seizures and mortality, blue states are Southern states with low and increasing seizures and mortality, and Western states are purple with lower and steady seizures and mortality. Data from NCHS and NFLIS.

Fentanyl seizures are strongly associated with fentanyl mortality. This is consistent with fentanyl as the cause of the third wave of the epidemic and these deaths (Zoorob 2019; O’Donnell et al. 2017; Hedegaard, Minino, and Warner 2018). Heroin seizures are also strongly associated with heroin mortality, although less so. Opioid seizures were least associated with opioid mortality, reflecting the fact that while illicit fentanyl availability is the main driver of fentanyl mortality, illicit opioid availability is not the only driver of all opioid mortality, as prescription opioids play a large role (Ahmad, Rossen, and Sutton 2020).

To evaluate the relationship between opioid seizures and mortality, we regressed fentanyl, heroin, and all opioid seizure rates respectively against fentanyl, heroin, and all opioid mortality on the log scale, while controlling for state and year fixed effects. The results are shown in Table 1 below. All associations were significant, and followed the patterns described above.

**Table 1:** Results from regression of different opioid seizure rates from NFLIS data against opioid mortality rates from NCHS all-cause mortality data, controlling for state and fixed effects.

| Opioid Type | Regression Coefficient of Standardized Seizures | Standard Error |
| --- | --- | --- |
| Heroin | 0.0491691 | 0.0330669 |
| Fentanyl | 0.5286017 | 0.0207397 |
| All Opioids | 0.0830363 | 0.0355304 |

Fentanyl seizures, heroin seizures, and opioid mortality also varied together by region across the US. In Figure 7, we plotted standardized fentanyl and heroin seizures alongside age-standardized opioid-related deaths. Northeastern, Midwestern and Appalachian states had high opioid mortality, high and increasing fentanyl seizure rates, and high heroin seizure rates. Southern states had low but increasing mortality and seizures, and Western states had lower and steady mortality and seizures.

## 5 Discussion

Opioid seizures, heroin seizures, and fentanyl seizures all varied by region, but had similar patterns within regions of the US. Seizures of all subtypes were high in Northeast, but remained relatively constant in the rest of the country. Ohio and Massachusetts had both the highest seizure counts and rates for heroin and fentanyl throughout the study period. In contrast to heroin and fentanyl, other opioid seizures didn’t follow regional patterns, and were decreasing, meaning national increases in opioid seizure rates are entirely due to increases in heroin and fentanyl seizures. The current literature suggests that heroin purity increased and price decreased in the late 2000s across the United States, because of a change in heroin origin and supply networks. Regional supply network changes could also explain this regional variation in fentanyl and heroin seizure rates.

Seizures of fentanyl and heroin together increased in certain states. Studies have implicated fentanyl-adulterated heroin or fentanyl sold as heroin in opioid mortality (Ciccarone 2017; Carroll et al. 2017; Kenney et al. 2018). This increase may indicate that fentanyl-adulterated heroin is present in the drug supply, or it could be due only to the increased presence of fentanyl in the drug supply, rather than fentanyl-adulteration. This dataset does not report fentanyl-adulterated heroin specifically, and does not indicate if fentanyl and heroin were found mixed together – only if they were seized together.

There were strong associations between drug seizures reported in the NFLIS dataset and drug-related mortality. Fentanyl seizures, heroin seizures, and all opioid seizures are associated with fentanyl, heroin, and all opioid deaths respectively. These results support existing literature claiming that the illicit drug supply is the main driver of the fentanyl epidemic, and that illicit supply played a role in heroin deaths from 2011 onward (Hedegaard, Minino, and Warner 2018; Kanouse and Compton 2015; Maxwell 2011; Mars, Rosenblum, and Ciccarone 2019). These results also suggest that there are regional patterns of drug supply and associated mortality, supporting existing literature suggesting regional drug supply changes influence the risks of drug use, in turn shaping overdose rates (Trinidad et al. 2016; Shiels et al. 2019; Zoorob 2019; O’Donnell et al. 2017; Hedegaard, Minino, and Warner 2018).

However, the NFLIS dataset is not a perfect descriptor of illicit drug supply or availability. Seizure reports are not random, and are subject to decisions about policing that may make certain drugs more likely to be seized, or communities differently policed, producing non-random patterns in seizure data. The dataset is therefore non-representative. Despite NLFIS covering 98% of analyses nation-wide, state lab systems and participating private labs represent only a percentage of the forensic analyses carried out in a state. In some states, large portions of analyses may be carried out in non-reporting private labs and be missing. Private labs may also analyze certain drugs more often than others (ex. all suspected heroin and no cannabis) biasing the NFLIS dataset to include fewer of those seizures. Unfortunately, we could not obtain state-specific coverage information for any states, so correcting for coverage differences or selection is impossible.

Additionally, some state lab systems may have changed how they analyze or report seizures during the study period, though NFLIS itself did not. Rosenblum et al. note in their 2021 paper that Ohio stopped testing for misdemeanor amounts of Cannabis in 2016, lowering the number of Cannabis reports recorded. Although we do not analyze Cannabis in this paper, similar changes may affect opioid reporting. As well, if the average quantity of drugs seized in a single report differs by state, or there are significantly more undercover buys in some states relative to seizures, artificially inflating the number of reports in the dataset relative to illicit drug supply, this could introduce noise.

Again, the existing literature suggests that illicit drug supply plays a large role in opioid mortality (Dasgupta, Beletsky, and Ciccarone 2018; Ruhm 2019). Therefore, despite these limitations, this literature along with the associations between seizures and mortality found here suggests that these data may be valuable in predicting drug mortality. This dataset is also the only national-level information available to researchers on national illicit drug supply. Though many improvements in data collection and quality would be necessary if this seizure data were to be trusted for information on drug supply or availability, or if it were to function as an early warning surveillance system for epidemics of illicit drug use, it has potential. However, if law enforcement data are improved for epidemiological surveillance, the resulting information must not used to maintain unjust law enforcement practices or support them.

The strong associations between seizures and mortality exhibited here mean these data show promise, and other seizure data show even more potential to predict drug overdoses (Rosenblum, Unick, and Ciccarone 2020a).

## Data Availability

All data produced are available online at https://github.com/heathermcb/national_seizure.

### Appendix

#### Categorization of opioids in NFLIS

The following is a table of the 100 most frequently listed drugs in the NFLIS dataset, along with our categorizations of these substances as described in ‘Methods’.

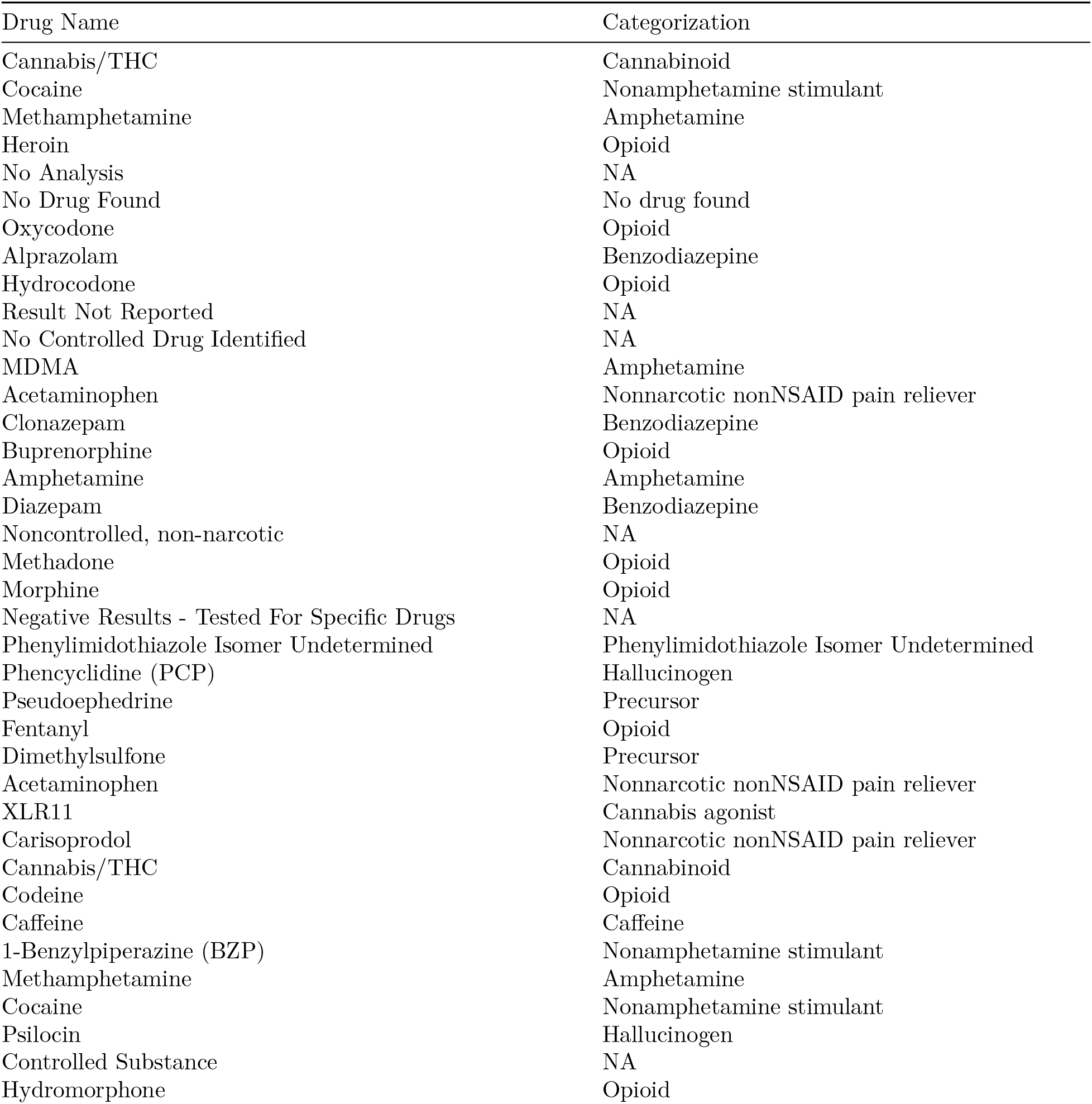
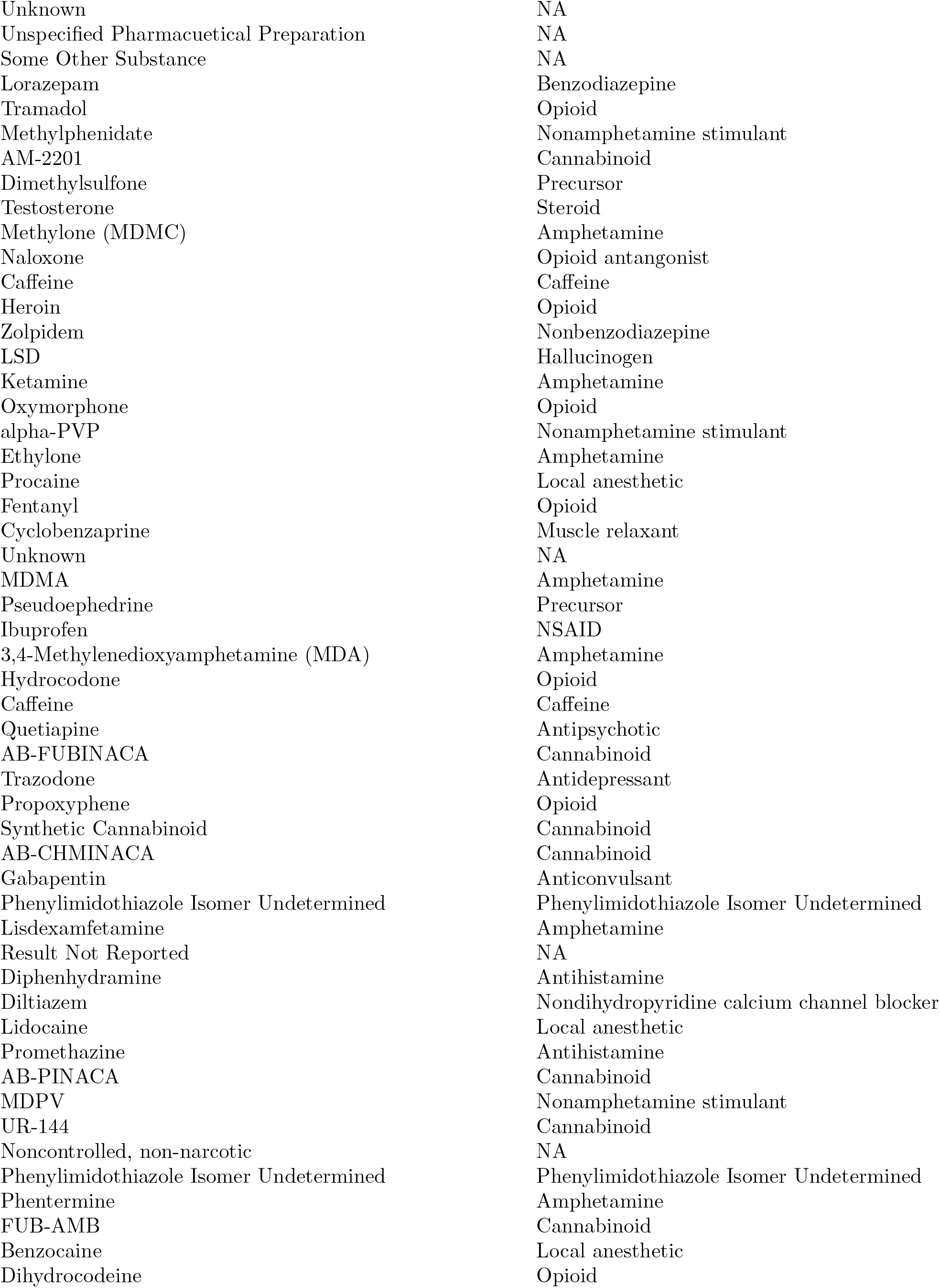
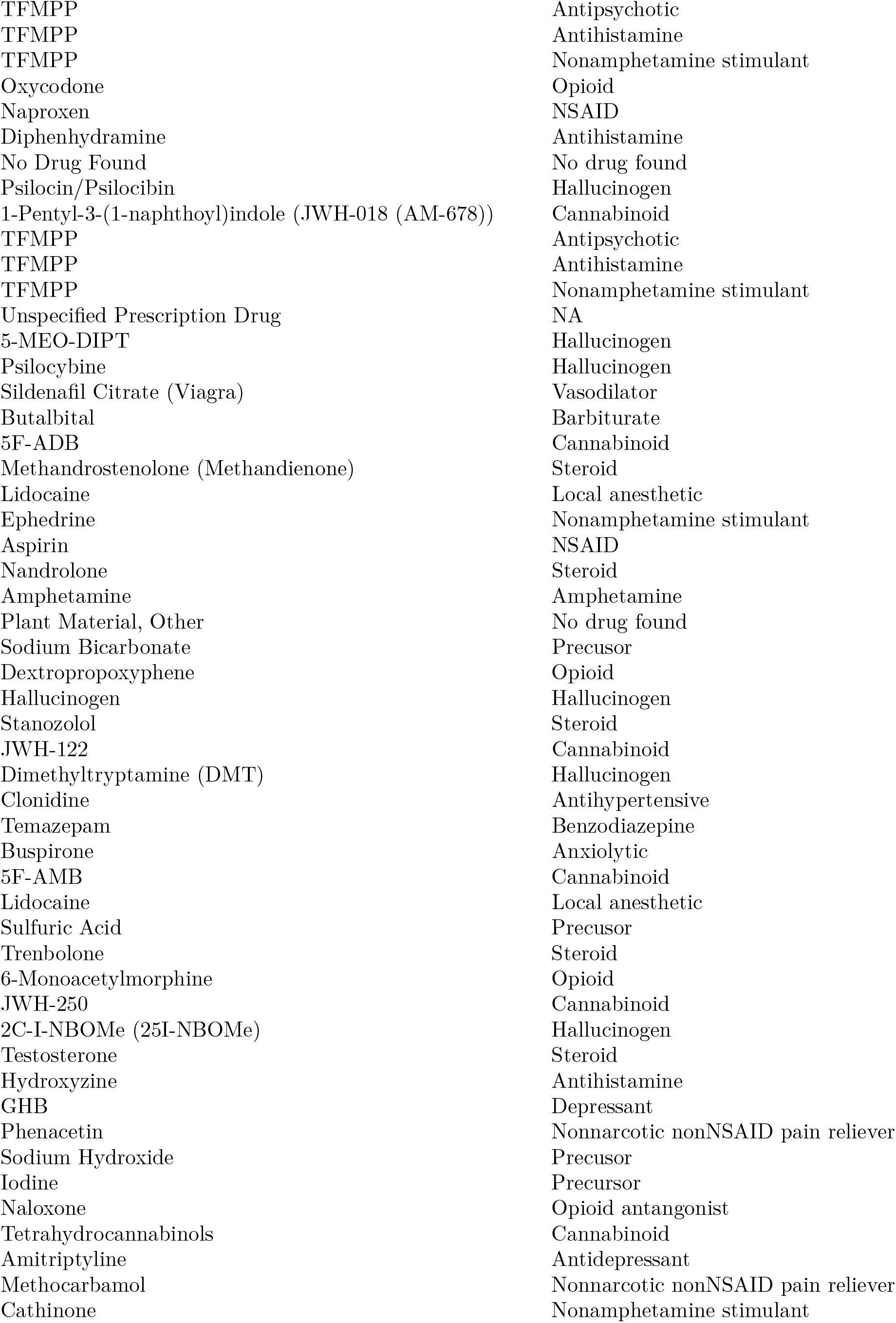
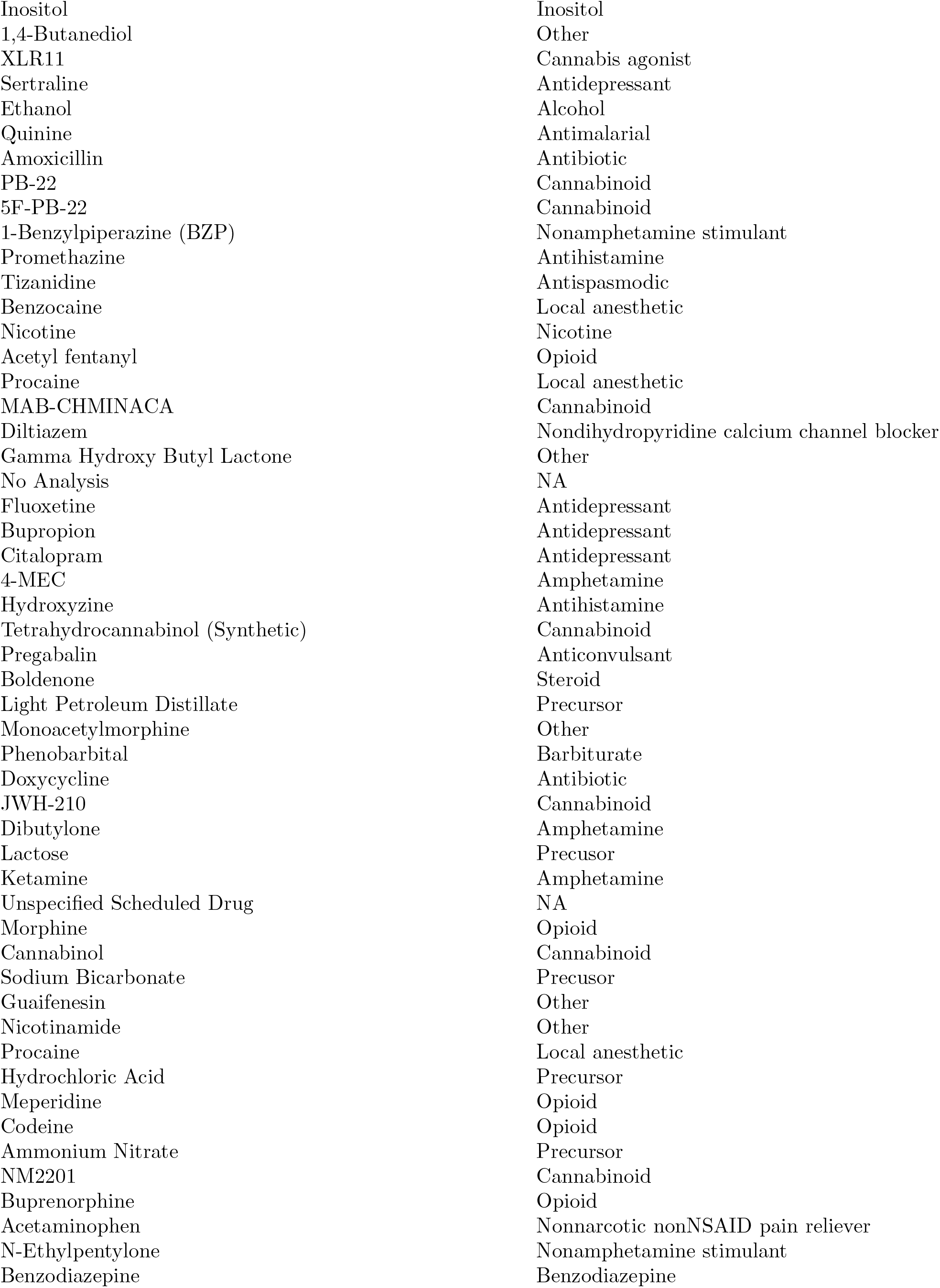
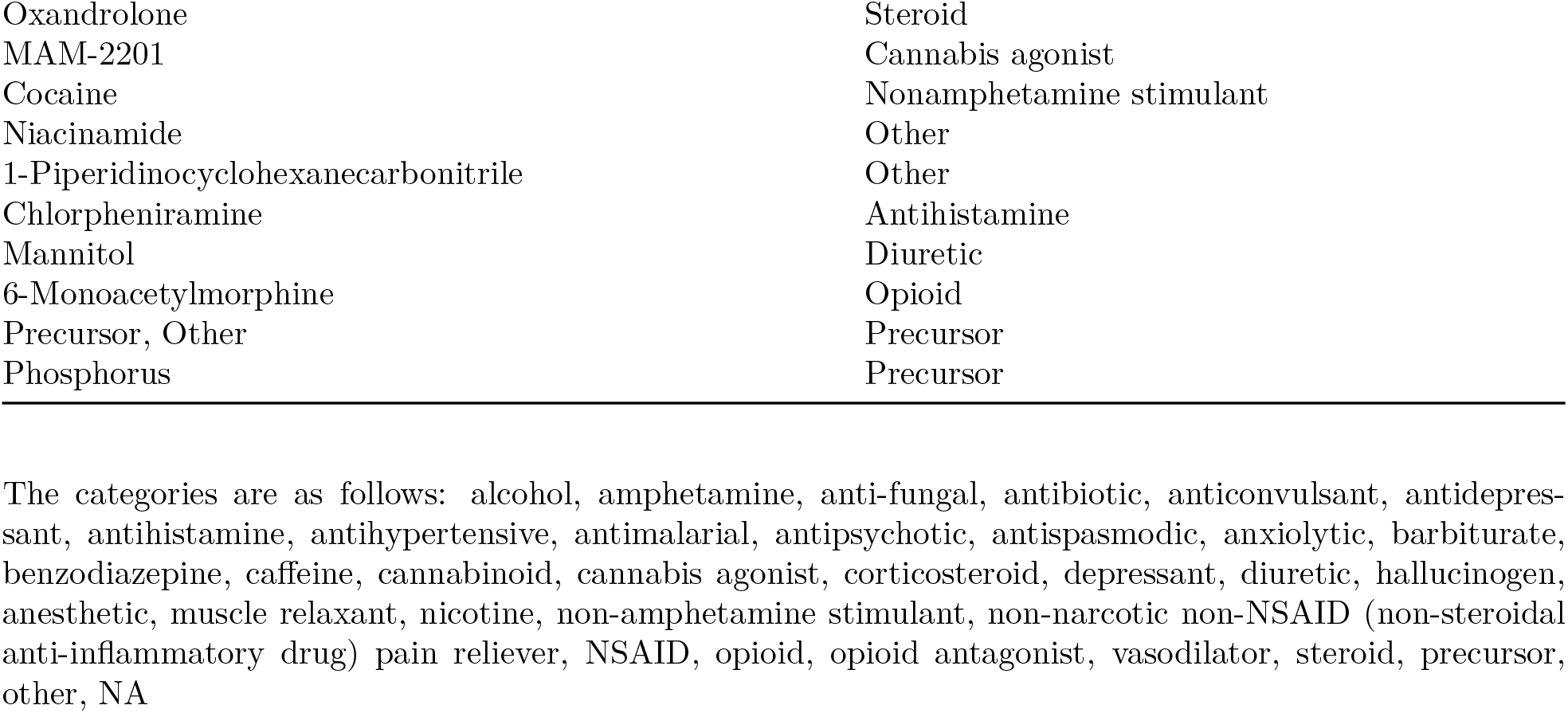

| Drug Name | Categorization |
| --- | --- |
| Cannabis/THC | Cannabinoid |
| Cocaine | Nonamphetamine stimulant |
| Methamphetamine | Amphetamine |
| Heroin | Opioid |
| No Analysis | NA |
| No Drug Found | No drug found |
| Oxycodone | Opioid |
| Alprazolam | Benzodiazepine |
| Hydrocodone | Opioid |
| Result Not Reported | NA |
| No Controlled Drug Identified | NA |
| MDMA | Amphetamine |
| Acetaminophen | Nonnarcotic nonNSAID pain reliever |
| Clonazepam | Benzodiazepine |
| Buprenorphine | Opioid |
| Amphetamine | Amphetamine |
| Diazepam | Benzodiazepine |
| Noncontrolled, non-narcotic | NA |
| Methadone | Opioid |
| Morphine | Opioid |
| Negative Results - Tested For Specific Drugs | NA |
| Phenylimidothiazole Isomer Undetermined | Phenylimidothiazole Isomer Undetermined |
| Phencyclidine (PCP) | Hallucinogen |
| Pseudoephedrine | Precursor |
| Fentanyl | Opioid |
| Dimethylsulfone | Precursor |
| Acetaminophen | Nonnarcotic nonNSAID pain reliever |
| XLR11 | Cannabis agonist |
| Carisoprodol | Nonnarcotic nonNSAID pain reliever |
| Cannabis/THC | Cannabinoid |
| Codeine | Opioid |
| Caffeine | Caffeine |
| 1-Benzylpiperazine (BZP) | Nonamphetamine stimulant |
| Methamphetamine | Amphetamine |
| Cocaine | Nonamphetamine stimulant |
| Psilocin | Hallucinogen |
| Controlled Substance | NA |
| Hydromorphone | Opioid |
| Unknown | NA |
| Unspecified Pharmacuetical Preparation | NA |
| Some Other Substance | NA |
| Lorazepam | Benzodiazepine |
| Tramadol | Opioid |
| Methylphenidate | Nonamphetamine stimulant |
| AM-2201 | Cannabinoid |
| Dimethylsulfone | Precursor |
| Testosterone | Steroid |
| Methylone (MDMC) | Amphetamine |
| Naloxone | Opioid antagonist |
| Caffeine | Caffeine |
| Heroin | Opioid |
| Zolpidem | Nonbenzodiazepine |
| LSD | Hallucinogen |
| Ketamine | Amphetamine |
| Oxymorphone | Opioid |
| alpha-PVP | Nonamphetamine stimulant |
| Ethylone | Amphetamine |
| Procaine | Local anesthetic |
| Fentanyl | Opioid |
| Cyclobenzaprine | Muscle relaxant |
| Unknown | NA |
| MDMA | Amphetamine |
| Pseudoephedrine | Precursor |
| Ibuprofen | NSAID |
| 3,4-Methylenedioxyamphetamine (MDA) | Amphetamine |
| Hydrocodone | Opioid |
| Caffeine | Caffeine |
| Quetiapine | Antipsychotic |
| AB-FUBINACA | Cannabinoid |
| Trazodone | Antidepressant |
| Propoxyphene | Opioid |
| Synthetic Cannabinoid | Cannabinoid |
| AB-CHMINACA | Cannabinoid |
| Gabapentin | Anticonvulsant |
| Phenylimidothiazole Isomer Undetermined | Phenylimidothiazole Isomer Undetermined |
| Lisdexamfetamine | Amphetamine |
| Result Not Reported | NA |
| Diphenhydramine | Antihistamine |
| Diltiazem | Nondihydropyridine calcium channel blocker |
| Lidocaine | Local anesthetic |
| Promethazine | Antihistamine |
| AB-PINACA | Cannabinoid |
| MDPV | Nonamphetamine stimulant |
| UR-144 | Cannabinoid |
| Noncontrolled, non-narcotic | NA |
| Phenylimidothiazole Isomer Undetermined | Phenylimidothiazole Isomer Undetermined |
| Phentermine | Amphetamine |
| FUB-AMB | Cannabinoid |
| Benzocaine | Local anesthetic |
| Dihydrocodeine | Opioid |
| TFMPP | Antipsychotic |
| TFMPP | Antihistamine |
| TFMPP | Nonamphetamine stimulant |
| Oxycodone | Opioid |
| Naproxen | NSAID |
| Diphenhydramine | Antihistamine |
| No Drug Found | No drug found |
| Psilocin/Psilocibin | Hallucinogen |
| 1-Pentyl-3-(1-naphthoyl)indole (JWH-018 (AM-678)) | Cannabinoid |
| TFMPP | Antipsychotic |
| TFMPP | Antihistamine |
| TFMPP | Nonamphetamine stimulant |
| Unspecified Prescription Drug | NA |
| 5-MEO-DIPT | Hallucinogen |
| Psilocybine | Hallucinogen |
| Sildenafil Citrate (Viagra) | Vasodilator |
| Butalbital | Barbiturate |
| 5F-ADB | Cannabinoid |
| Methandrostenolone (Methandienone) | Steroid |
| Lidocaine | Local anesthetic |
| Ephedrine | Nonamphetamine stimulant |
| Aspirin | NSAID |
| Nandrolone | Steroid |
| Amphetamine | Amphetamine |
| Plant Material, Other | No drug found |
| Sodium Bicarbonate | Precursor |
| Dextropropoxyphene | Opioid |
| Hallucinogen | Hallucinogen |
| Stanozolol | Steroid |
| JWH-122 | Cannabinoid |
| Dimethyltryptamine (DMT) | Hallucinogen |
| Clonidine | Antihypertensive |
| Temazepam | Benzodiazepine |
| Buspirone | Anxiolytic |
| 5F-AMB | Cannabinoid |
| Lidocaine | Local anesthetic |
| Sulfuric Acid | Precursor |
| Trenbolone | Steroid |
| 6-Monoacetylmorphine | Opioid |
| JWH-250 | Cannabinoid |
| 2C-I-NBOMe (25I-NBOMe) | Hallucinogen |
| Testosterone | Steroid |
| Hydroxyzine | Antihistamine |
| GHB | Depressant |
| Phenacetin | Nonnarcotic nonNSAID pain reliever |
| Sodium Hydroxide | Precursor |
| Iodine | Precursor |
| Naloxone | Opioid antagonist |
| Tetrahydrocannabinols | Cannabinoid |
| Amitriptyline | Antidepressant |
| Methocarbamol | Nonnarcotic nonNSAID pain reliever |
| Cathinone | Nonamphetamine stimulant |
| Inositol | Inositol |
| 1,4-Butanediol | Other |
| XLR11 | Cannabis agonist |
| Sertraline | Antidepressant |
| Ethanol | Alcohol |
| Quinine | Antimalarial |
| Amoxicillin | Antibiotic |
| PB-22 | Cannabinoid |
| 5F-PB-22 | Cannabinoid |
| 1-Benzylpiperazine (BZP) | Nonamphetamine stimulant |
| Promethazine | Antihistamine |
| Tizanidine | Antispasmodic |
| Benzocaine | Local anesthetic |
| Nicotine | Nicotine |
| Acetyl fentanyl | Opioid |
| Procaine | Local anesthetic |
| MAB-CHMINACA | Cannabinoid |
| Diltiazem | Nondihydropyridine calcium channel blocker |
| Gamma Hydroxy Butyl Lactone | Other |
| No Analysis | NA |
| Fluoxetine | Antidepressant |
| Bupropion | Antidepressant |
| Citalopram | Antidepressant |
| 4-MEC | Amphetamine |
| Hydroxyzine | Antihistamine |
| Tetrahydrocannabinol (Synthetic) | Cannabinoid |
| Pregabalin | Anticonvulsant |
| Boldenone | Steroid |
| Light Petroleum Distillate | Precursor |
| Monoacetylmorphine | Other |
| Phenobarbital | Barbiturate |
| Doxycycline | Antibiotic |
| JWH-210 | Cannabinoid |
| Dibutylone | Amphetamine |
| Lactose | Precursor |
| Ketamine | Amphetamine |
| Unspecified Scheduled Drug | NA |
| Morphine | Opioid |
| Cannabinol | Cannabinoid |
| Sodium Bicarbonate | Precursor |
| Guaifenesin | Other |
| Nicotinamide | Other |
| Procaine | Local anesthetic |
| Hydrochloric Acid | Precursor |
| Meperidine | Opioid |
| Codeine | Opioid |
| Ammonium Nitrate | Precursor |
| NM2201 | Cannabinoid |
| Buprenorphine | Opioid |
| Acetaminophen | Nonnarcotic nonNSAID pain reliever |
| N-Ethylpentylone | Nonamphetamine stimulant |
| Benzodiazepine | Benzodiazepine |
| Oxandrolone | Steroid |
| MAM-2201 | Cannabis agonist |
| Cocaine | Nonamphetamine stimulant |
| Niacinamide | Other |
| 1-Piperidinocyclohexanecarbonitrile | Other |
| Chlorpheniramine | Antihistamine |
| Mannitol | Diuretic |
| 6-Monoacetylmorphine | Opioid |
| Precursor, Other | Precursor |
| Phosphorus | Precursor |

The categories are as follows: alcohol, amphetamine, anti-fungal, antibiotic, anticonvulsant, antidepressant, antihistamine, antihypertensive, antimalarial, antipsychotic, antispasmodic, anxiolytic, barbiturate, benzodiazepine, caffeine, cannabinoid, cannabis agonist, corticosteroid, depressant, diuretic, hallucinogen, anesthetic, muscle relaxant, nicotine, non-amphetamine stimulant, non-narcotic non-NSAID (non-steroidal anti-inflammatory drug) pain reliever, NSAID, opioid, opioid antagonist, vasodilator, steroid, precursor, other, NA

## References

Ahmad, FB, LM Rossen, and P Sutton. 2020. “Provisional Drug Overdose Death Counts.” National Center for Health Statistics. https://www.cdc.gov/nchs/nvss/vsrr/drug-overdose-data.htm.

Beletsky, Leo, and Corey S. Davis. 2017. “Today’s Fentanyl Crisis: Prohibition’s Iron Law, Revisited.” International Journal of Drug Policy 46: 156–59. https://doi.org/10.1016/j.drugpo.2017.05.050.

Bohnert, Amy, Marcia Valenstein, Matthew Bair, and et al. 2011. “Association Between Opioid Prescribing Patterns and Opioid Overdose-Related Deaths.” JAMA 305 (13): 1315–21. https://jamanetwork.com/journals/jama/fullarticle/896182.

Carroll, Jennifer J., Brandon D. L. Marshall, Josiah D. Rich, and Traci C. Green. 2017. “Exposure to Fentanyl-Contaminated Heroin and Overdose Risk Among Illicit Opioid Users in Rhode Island: A Mixed Methods Study.” International Journal of Drug Policy 46: 136–45. https://doi.org/10.1016/j.drugpo.2017.05.023.

Chou, Roger, Judith Turner, Emily Devine, Richard Deyo, and et al. 2015. “The Effectiveness and Risks of Long-Term Opioid Therapy for Chronic Pain: A Systematic Review for a National Institutes of Health Pathways to Prevention Workshop.” Annals of Internal Medicine 162 (4): 276–86. https://www.acpjournals.org/doi/abs/10.7326/M14-2559.

Ciccarone, Daniel. 2017. “Fentanyl in the US Heroin Supply: A Rapidly Changing Risk Environment.” International Journal of Drug Policy 46: 107–11. https://doi.org/10.1016/j.drugpo.2017.06.010.

Ciccarone, Daniel. 2019. “The Triple Wave Epidemic: Supply and Demand Drivers of the US Opioid Overdose Crisis.” International Journal on Drug Policy.

Ciccarone, Daniel, Jeff Ondocsin, and Sarah G. Mars. 2017. “Heroin Uncertainties: Exploring Users’ Perceptions of Fentanyl-Adulterated and -Substituted ‘Heroin’.” International Journal of Drug Policy 46: 146–55. https://doi.org/10.1016/j.drugpo.2017.06.004.

Cicero, Theodore, Matthew Ellis, Hilary Surratt, and Steven Kurtz. 2014. “The Changing Face of Heroin Use in the United States: A Retrospective Analysis of the Past 50 Years.” Jama Psychiatry 71 (7): 821–26. https://pubmed.ncbi.nlm.nih.gov/24871348/.

Dart, Richard C., Hilary L. Surratt, Theodore J. Cicero, Mark W. Parrino, S. Geoff Severtson, Becki Bucher-Bartelson, and Jody L. Green. 2015. “Trends in Opioid Analgesic Abuse and Mortality in the United States.” New England Journal of Medicine 372 (3): 241–48. https://doi.org/10.1056/NEJMsa1406143.

Dasgupta, Nabarun, Leo Beletsky, and Daniel Ciccarone. 2018. “Opioid Crisis: No Easy Fix to Its Social and Economic Determinants.” American Journal of Public Health 108 (2): 182–86. https://doi.org/10.2105/AJPH.2017.304187.

Drug Enforcement Administration. 2016. “National Heroin Threat Assessment Summary.” Department of Homeland Security. https://www.hsdl.org/?abstract&did=797265.

Drug Enforcement Administration. 2017. “2017 National Drug Threat Assessment.” Department of Homeland Security. https://www.dea.gov/sites/default/files/2018-07/DIR-040-17_2017-NDTA.pdf.

Drug Enforcement Administration. 2020. “2019 National Drug Threat Assessment.” Department of Homeland Security. https://www.dea.gov/documents/2020/01/30/2019-national-drug-threat-assessment.

Drugs, United Nations Office on, and Crime. 2012. “World Drug Report.” United Nations. https://www.unodc.org/documents/data-and-analysis/WDR2012/WDR_2012_web_small.pdf.

Fischer, B, M Pang, and W Jones. 2020. “The Opioid Mortality Epidemic in North America: Do We Understand the Supply Side Dynamics of This Unprecedented Crisis?” Subst Abuse Treat Prev Policy 15 (14). https://substanceabusepolicy.biomedcentral.com/articles/10.1186/s13011-020-0256-8#citeas.

Fulton-Kehoe, Deborah, Mark Sullivan, Judith Turner, Renu Garg, Amy Bauer, Thomas Wickizer, and Gary Franklin. 2015. “Opioid Poisonings in Washington State Medicaid.” Medical Care 53 (8): 679–85. https://journals.lww.com/lww-medicalcare/Abstract/2015/08000/Opioid_Poisonings_in_Washington_State_Medicaid_.4.aspx.

Haley, Danielle F., and Richard Saitz. 2020. “The Opioid Epidemic During the COVID-19 Pandemic.” JAMA 324 (16): 1615–17. https://jamanetwork.com/journals/jama/fullarticle/2770985.

Hedegaard, Holly, Arialdi M Minino, and Margaret Warner. 2018. “Drug Overdose Deaths in the United States, 1999–2017. NCHS Data Brief, No 329.” National Center for Health Statistics, Hyattsville, MD.

Kanouse, Andrew B, and Peggy Compton. 2015. “The Epidemic of Prescription Opioid Abuse, the Subsequent Rising Prevalence of Heroin Use, and the Federal Response.” Journal of Pain & Palliative Care Pharmacotherapy 29 (2): 102–14.

Kenney, Shannon R., Bradley J. Anderson, Micah T. Conti, Genie L. Bailey, and Michael D. Stein. 2018. “Expected and Actual Fentanyl Exposure Among Persons Seeking Opioid Withdrawal Management.” Journal of Substance Abuse Treatment 86: 65–69. https://doi.org/10.1016/j.jsat.2018.01.005.

Macmadu, Alexandria, Jennifer J. Carroll, Scott E. Hadland, Traci C. Green, and Brandon D. L. Marshall. 2017. “Prevalence and Correlates of Fentanyl-Contaminated Heroin Exposure Among Young Adults Who Use Prescription Opioids Non-Medically.” Addictive Behaviors 68: 35–38. https://doi.org/10.1016/j.addbeh.2017.01.014.

Mars, Sarah G., Jeff Ondocsin, and Daniel Ciccarone. 2018. “Sold as Heroin: Perceptions and Use of an Evolving Drug in Baltimore, MD.” Journal of Psychoactive Drugs 50 (2): 167–76. https://doi.org/10.1080/02791072.2017.1394508.

Mars, Sarah G., Daniel Rosenblum, and Daniel Ciccarone. 2019. “Illicit Fentanyls in the Opioid Street Market: Desired or Imposed?” Addiction 114 (5): 774–80. https://doi.org/10.1111/add.14474.

Maxwell, Jane Carlisle. 2011. “The Prescription Drug Epidemic in the United States: A Perfect Storm.” Drug and Alcohol Review 30 (3): 264–70.

Miller, Matthew, Catherine Barber, Sarah Leatherman, and et al. 2015. “Prescription Opioid Duration of Action and the Risk of Unintentional Overdose Among Patients Receiving Opioid Therapy.” JAMA Internal Medicine 174 (4): 608–15. https://jamanetwork.com/journals/jamainternalmedicine/fullarticle/2110997.

National Forensic Laboratory Information System. 2017. “Special Report: Opiates and Related Drugs Reported in NFLIS, 2009-2014.” Drug Enforcement Agency. https://www.nflis.deadiversion.usdoj.gov/DesktopModules/ReportDownloads/Reports/NFLIS-SR-Opioids-Rev-201702.pdf.

National Forensic Laboratory Information System. 2020a. “About NFLIS.” Drug Enforcement Agency. https://www.nflis.deadiversion.usdoj.gov/#:~:text=The%20National%20Forensic%20Laboratory%20Information,%2C%20regional%2C%20and%20national%20entities.

National Forensic Laboratory Information System. 2020b. “NFLIS 2020 Midyear Report.” https://www.nflis.deadiversion.usdoj.gov/nflisdata/docs/13915NFLISdrugMidYear2020.pdf.

National Forensic Laboratory Information System. 2020c. “NFLIS-Drug 2019 Annual Report.” Drug Enforcement Agency. https://www.nflis.deadiversion.usdoj.gov/DesktopModules/ReportDownloads/Reports/NFLIS-Drug-AR2019.pdf.

O’Donnell, Julie, John Halpin, Christine Mattson, Bruce Goldberger, and Matthew Gladden. 2017. “Deaths Involving Fentanyl, Fentanyl Analogs, and u-47700 — 10 States, July–December 2016.” Morbidity and Mortality Weekly Report (MMWR) 66 (43): 1197–1202. https://www.cdc.gov/mmwr/volumes/66/wr/mm6643e1.htm.

Paulozzi, Leonard, Christopher Jones, Karin Mack, and Rose Rudd. 2011. “Vital Signs: Overdoses of Prescription Opioid Pain Relievers – United States, 1999-2008.” Morbidity and Mortality Weekly Report (MMWR). https://www.cdc.gov/mmwr/preview/mmwrhtml/mm6043a4.htm.

Rosenblum, Daniel, Jay Unick, and Daniel Ciccarone. 2014. “The Entry of Colombian-Sourced Heroin into the US Market: The Relationship Between Competition, Price, and Purity.” International Journal of Drug Policy 25 (1): 88–95. https://www.ncbi.nlm.nih.gov/pmc/articles/PMC3946788/.

Rosenblum, Daniel, Jay Unick, and Daniel Ciccarone. 2020a. “The Rapidly Changing US Illicit Drug Market and the Potential for an Improved Early Warning System: Evidence from Ohio Drug Crime Labs.” Drug and Alcohol Dependence 208: 107779.

Rosenblum, Daniel, Jay Unick, and Daniel Ciccarone. 2020b. “The Rapidly Changing US Illicit Drug Market and the Potential for an Improved Early Warning System: Evidence from Ohio Drug Crime Labs.” Drug and Alcohol Dependence. https://www.sciencedirect.com/science/article/abs/pii/S0376871619305563?via%3Dihub.

Ruhm, Christopher J. 2019. “Drivers of the Fatal Drug Epidemic.” Journal of Health Economics 64: 25–42.

Shiels, Meredith S, Amy Berrington de González, Ana F Best, Yingxi Chen, Pavel Chernyavskiy, Patricia Hartge, Sahar Q Khan, et al. 2019. “Premature Mortality from All Causes and Drug Poisonings in the USA According to Socioeconomic Status and Rurality: An Analysis of Death Certificate Data by County from 2000–15.” The Lancet Public Health 4 (2): e97–106.

Trinidad, James P, Margaret Warner, BS Bastian, Arialdi M Miniño, Holly Hedegaard, et al. 2016. “National Vital Statistics Reports.” National Vital Statistics Reports 65 (9).

Woolf, SH., and H. Schoomaker. 2019. “Life Expectancy and Mortality Rates in the United States, 1959-2017.” JAMA 332 (20): 1935–2032. https://jamanetwork.com/journals/jama/issue/322/20.

Zoorob, Michael. 2019. “Fentanyl Shock: The Changing Geography of Overdose in the United States.” International Journal of Drug Policy 70: 40–46. https://pubmed.ncbi.nlm.nih.gov/31079029/.

